# Assessing the impact of temperature and humidity exposures during early infection stages on case-fatality of COVID-19: a modelling study in Europe

**DOI:** 10.1101/2021.09.23.21264017

**Authors:** Jingbo Liang, Hsiang-Yu Yuan

**Affiliations:** Department of Biomedical Sciences, Jockey Club College of Veterinary Medicine and Life Sciences, City University of Hong Kong, Hong Kong; Centre for Applied One Health Research and Policy Advice, City University of Hong Kong, Hong Kong

**Keywords:** Case fatality rate, Temperature, Humidity, Symptom onset, Immunity response

## Abstract

**Background:** Although associations between key weather indicators (i.e. temperature and humidity) and COVID-19 mortality has been reported, the relationship between these exposures among different timing in early infection stages (from virus exposure up to a few days after symptom onset) and the probability of death after infection (also called case fatality rate, CFR) has yet to be determined.

**Methods:** We estimated the instantaneous CFR of eight European countries using Bayesian inference in conjunction with stochastic transmission models, taking account of delays in reporting the number of newly confirmed cases and deaths. The exposure-lag–response associations between fatality rate and weather conditions to which patients were exposed at different timing were obtained using distributed lag nonlinear models coupled with mixed-effect models.

**Results:** Our results showed that the Odds Ratio (OR) of death is negatively associated with the temperature, with two maxima (OR=1.29 (95% CI: 1.23, 1.35) at -0.1°C; OR=1.12 (95% CI: 1.08, 1.16) at 0.1°C) occurred at the time of virus exposure and after symptom onset. Two minima (OR=0.81 (95% CI: 0.71, 0.92) at 23.2°C; OR=0.71 (95% CI: 0.63, 0.80) at 21.7°C) also occurred at these two distinct periods correspondingly. Low humidity (below 50%) during the early stages and high humidity (approximately 89%) after symptom onset were related to the lower fatality.

**Conclusion:** Environmental conditions may affect not only the initial viral load when exposure to viruses but also individuals’ immunity response around symptom onset. Warmer temperatures and higher humidity after symptom onset were related to the lower fatality.

**Highlights:**

- Temperature and humidity conditions that patients were exposed to during their early infection stages were associated with COVID-19 case fatality rate.
- Warmer temperatures (> 20°C) at infection time or after symptom onset, but not during the incubation period, were associated with lower death risk. Low relative humidity (< 50%) during the early stages and high relative humidity (> 85%) after symptom onset were related to higher death risk.
- Creating optimal indoor conditions for cases who are under quarantine/isolation may reduce their risk of death.

## 1. Introduction

The Coronavirus Disease 2019 (COVID-19) pandemic, attributable to the severe acute respiratory syndrome coronavirus 2 (SARS-CoV-2), has posed unprecedented challenges to the world including many European countries. The number of deaths in Europe (1,116,017) counts for approximately 27% of all COVID-19 deaths worldwide (data accessed on July 17, 2021 [1]). The relationship between weather conditions and COVID-19 fatality rates in Europe have not been fully determined. Furthermore, the impact of the weather conditions to which patients were exposed during the early infection stages (i.e. between virus infection and a few days after symptom onset) is unknown.

Temperature and humidity can likely influence the outcome of infection in two ways: affecting the initial viral load and modulating the immune response in patients. A possible distinction between these two mechanisms is the time point at which environmental risk factors take effect during the course of infection. The stability and the viability of SARS-CoV-2 are likely to be affected by environmental conditions when individuals are exposed to the virus [2]. A higher initial viral load may pose a higher risk for developing severe diseases later. On the other hand, the local innate immune responses that occur in the upper or lower respiratory tract could be activated immediately after symptom onset [3]. The dynamics of the innate response can be influenced by temperature or humidity [4, 5], presumably, during the time when it was activated.

According to the states of immune responses and pathogenesis, the infection course of SARS-CoV-2 could be divided into three stages: asymptomatic incubation period, non-severe symptomatic period, and severe respiratory symptomatic stage [6]. If certain controllable environmental conditions (e.g. temperature and humidity) during the non-severe symptomatic period can affect patients’ innate immune responses [7, 8, 4] and hence their risk of death, preventive measures can be designed to reduce COVID-19 severity for these cases. However, until now, no such preventive measures are proposed due to that the impact of those conditions during the symptomatic period is unknown.

Although many studies have reported that low temperatures may increase the COVID-19 deaths or mortality [9, 10, 11, 12, 13, 14], these studies did not have a direct measurement on the risk of death. Case fatality rate (CFR) [15, 16] is an important index to measure the disease severity, but one limitation is that this rate only represents the average proportion of deaths among all confirmed cases over a duration of time, without the ability to reflect the instantaneous probability of death. This time-varying instantaneous probability, also called instantaneous CFR (iCFR) [17], can be influenced by many factors, such as change in health care capacity [18] and variations in weather conditions [19, 20], etc. Challenges exist in estimating the iCFR due to the time delays between confirming a positive case and reporting his/her outcome [15, 21]. A more accurate estimation of the iCFR can be obtained if these delays are incorporated in modelling.

The study aimed at assessing the impact of weather conditions COVID-19 patients were exposed to at different timing of the early infection course on the death risk. To resolve the above issues in delays and to estimate the iCFR, stochastic modelling [22] was used, taking into account of the delays in reporting the number of newly confirmed cases and deaths in each of the European countries. After adjusting delays in reporting cases and deaths, the correlation between the iCFR and daily weather conditions at different timing since the infection was obtained using distributed lag non-linear models (DLNMs) coupled with generalized linear mixed models.

## 2. Material and Methods

### 2.1. Data collection

Our study focused on eight European countries (the United Kingdom, Italy, France, Spain, Germany, Netherlands, Sweden, and Romania) where the cumulative number of deaths was larger than 2000 during the first wave of the pandemic. The daily numbers of reported COVID-19 cases and deaths in these eight European countries from 16th February to 31st June 2020 were collected from ‘Our World in Data’ [1]. We defined the community outbreak began from the first two consecutive days whose average case number exceeded the country’s baseline (the 5% quantile of the maximal daily number of new cases) and ended at the first two consecutive days whose average case number was less than that baseline.

Daily mean temperatures for the eight European countries and daily relative humidity values for five countries (Italy, Spain, Germany, Netherlands, and Sweden) were collected from the European Climate Assessment and Dataset (ECAD)[23]. For countries (the United Kindom, France, and Romania) lacking relative humidity data, we calculated relative humidity values using the ratio of the actual water vapor pressure divided by the saturation water vapor pressure[13] (see Supplementary Methods). In this study, the daily mean temperatures and relative humidity values for each country were calculated using the average records from all monitoring stations.

### 2.2. Estimating COVID-19 transmission patterns by SEIR model

We constructed a stochastic model that extended Susceptible - Exposed - Infectious - Recovered (SEIR) model to reproduce the COVID-19 transmission dynamic and estimate the iCFR and the effective reproduction number (Re) for each of countries during their outbreaks. The extended SEIR model contained three additional compartments: HR, hospital confirmed cases who later recovered; HD, hospital confirmed cases who later died of infection; and D, total deaths (Figure 1). In order to calculate the iCFR of the date of case confirmation, newly confirmed cases were divided into HR and HD compartments following the probabilities of 1-iCFR and iCFR upon the date of case confirmation. Delays for case confirmation and death reporting were included. To estimate parameters in stochastic models, the Particle Markov chain Monte Carlo (PMCMC) method [22, 24], a combination of particle filtering and Markov chain Monte Carlo approaches was used (See Supplementary Methods). Posterior distributions of all parameters used in the model were obtained using the Nimble package in R [25] (version 3.6.1). The settings of prior distributions for these parameters were provided in Supplementary Methods.

**Figure 1:**
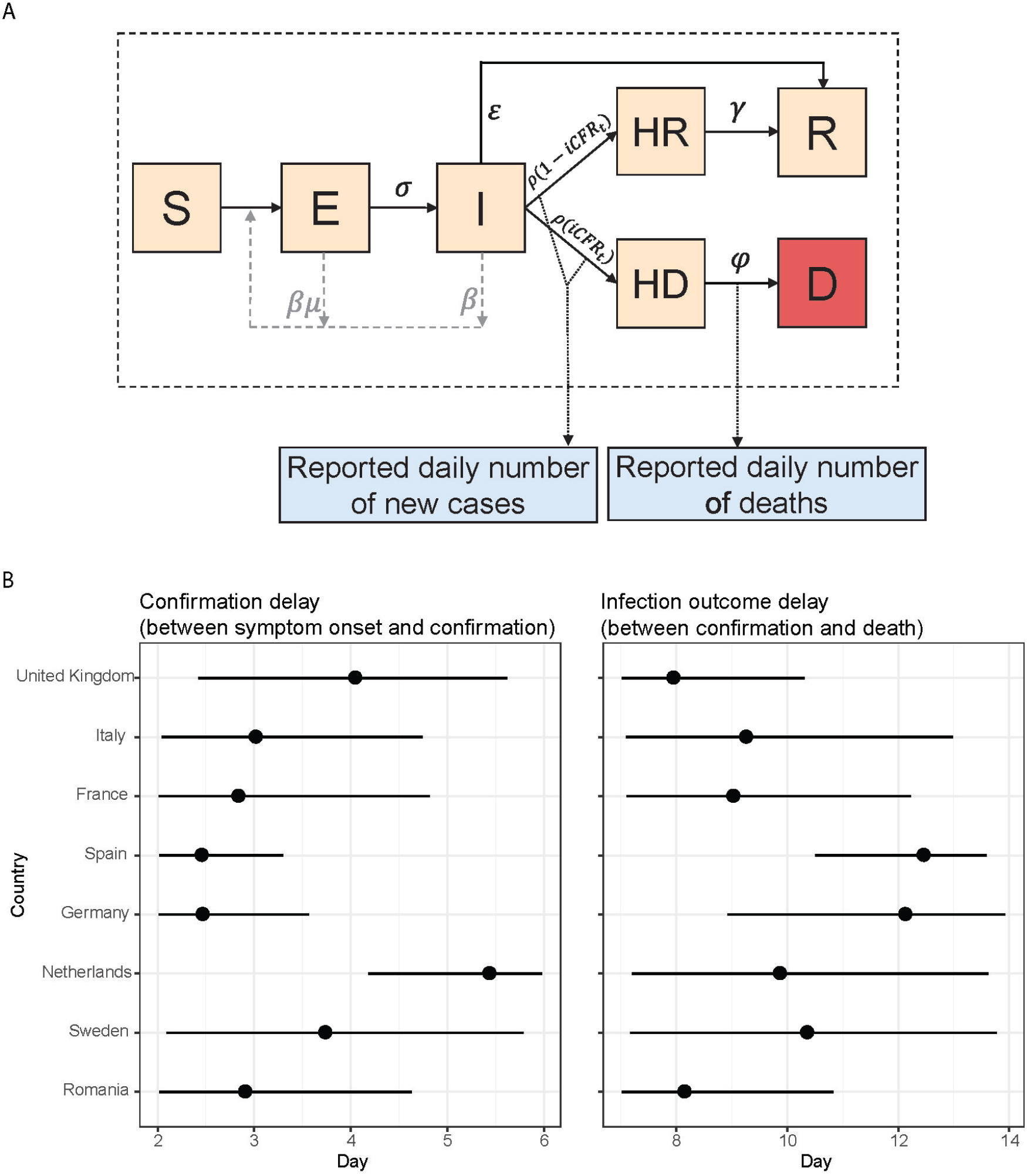
(A) Schematic of the extended Susceptible, Exposed, Infectious and Recovered model with case confirmation and death statuses. The total population was divided into seven compartments: S (susceptible), E (exposed), I (infectious after the incubation period), HR (hospital confirmed cases who later recovered), HD (hospital confirmed cases who later died), R (recovered), and D (death). *β* is the transmission rate, 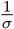 is the incubation period, *μ* is the proportion of pre-symptomatic infectious individuals among the total number of exposed individuals [44], *ε* is the recovery rate for un-reported cases (mainly asymptomatic cases), 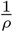 is the confirmation delay, 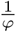 is the infection outcome delay, and *γ* is the recovery rate for hospital confirmed cases. *iCFR*_*t*_ is the iCFR at time *t*. (B) Estimated confirmation delay and infection outcome delay in the eight European countries. Black dots represent the posterior mean value and horizontal lines represent the 95% posterior credible intervals.

### 2.3. Estimating the effects of temperature and relative humidity on the COVID-19 iCFR based on a DLNM model

In order to avoid the impacts from the variations in non-pharmaceutical interventions (NPIs) [26], we estimated the effects of weather conditions during the epidemic period when Re remained relatively stable below 1.5. Presumably, the variations of NPIs were assumed to be minor during this period.

To explore the non-linear association between weather conditions and the iCFR with taking account of other unknown local factors for each of countries, a combination of distributed lag nonlinear models (DLNMs) [27] and generalized linear mixed models (GLMM) was adopted to estimate the differential effects of weather conditions exposed during the early infection stages on the iCFR. For detailed steps, please see Supplementary Methods.

### 2.4. Model validation

To assess convergence of parameters in the SEIR model, we constructed three independent chains of algorithm sets with 100,000 iterations and calculated the Gelman-Rubin convergence diagnostic statistics [28] across the three chains. For DLNM models, we used different combinations of temperature and relative humidity as predictors. The model with the lowest Akaike information criterion (AIC) and Bayesian information criterion (BIC) was chosen as the best-fitting model. The prediction performance of the best-fitting model was presented by comparing its model-predicted iCFR with the iCFR calculated by SEIR models (Supplementary Figure 1).

## 3. Results

### 3.1. Case fatality in Europe

In order to estimate the iCFR among the eight European countries, we developed a stochastic epidemic model (See Method and Figure 1A) taking account of delays between symptom onset and the confirmation of infection (i.e. confirmation delay) and delays between the confirmation and death (i.e. infection outcome delay). The model estimated the mean confirmation delays varied from 2.4 to 5.4 days, and the mean infection outcome delays varied from 7.9 to 12.4 days among these countries (Figure 1B, Supplementary Table 1). Incorporating the variations of such delays allowed a more accurate estimation of the country-specific iCFR and hence its relationship with weather conditions.

Our model successfully captured the dynamics of the daily new cases and deaths during the first wave (Figure 2, Supplementary Table 2). The epidemic patterns were similar (reaching a maximum of approximately or over eighty cases per one million people per day in April) among most of the countries, except Sweden, and Romania. Generally, the daily maximum number of deaths occurred in April. Four countries (including the United Kingdom, Italy, France, and Spain; see Figure 2A) had higher mortality rates (number of deaths per one million people): Spain had the highest daily estimated mortality rate of 18, followed by the United Kingdom 14, Italy 13, and France 11. The remaining countries (including Germany, Netherlands, Sweden and Romania; see Figure 2B) demonstrated lower mortality rates during similar periods: Netherlands had the highest daily estimated mortality rate of 9, followed by Sweden 7, Germany 3 and Romania 2.

**Figure 2:**
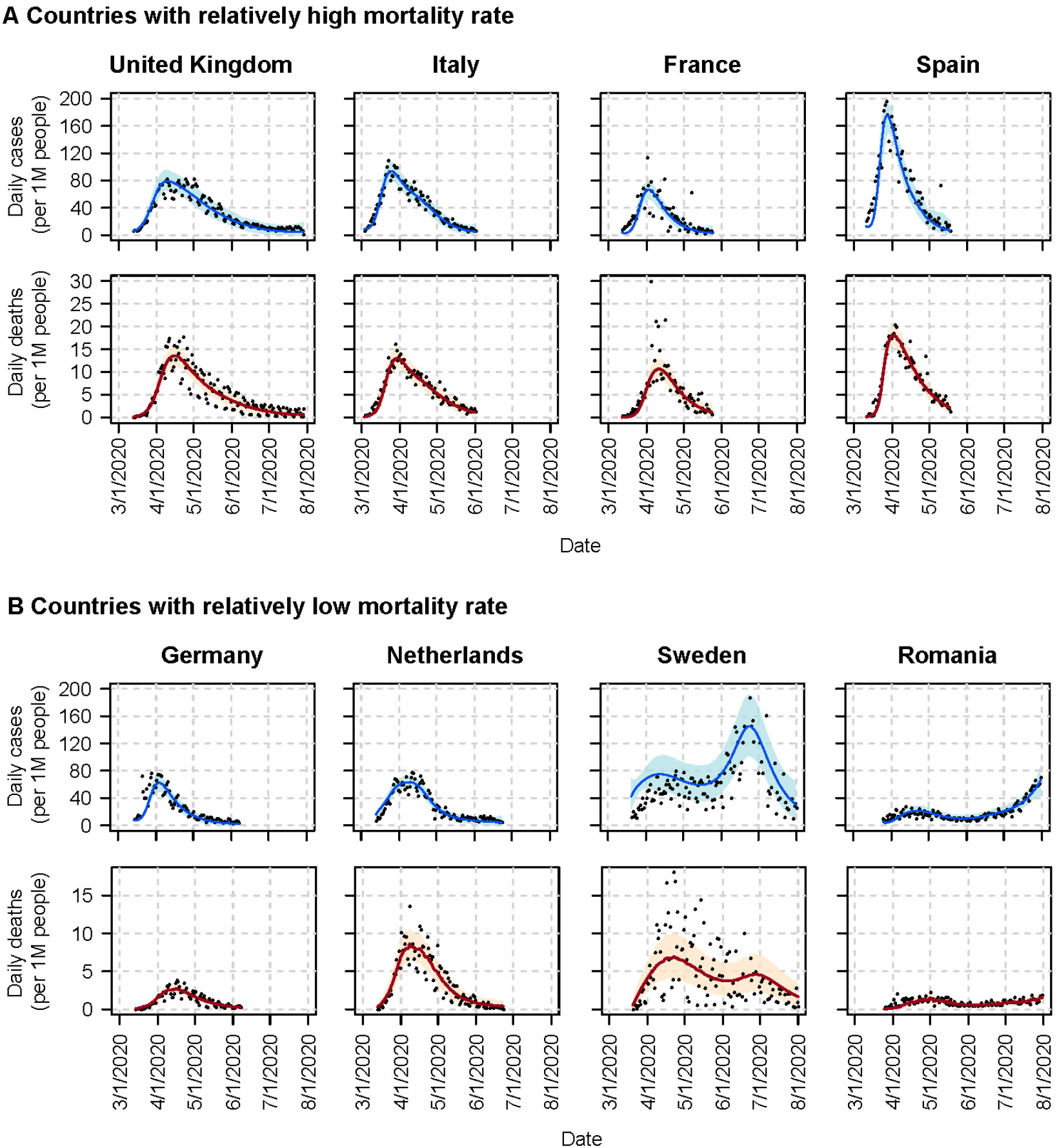
Observed and model-estimated daily numbers of cases and deaths per one million people in the eight European countries during the outbreaks. (A) Daily number of cases and deaths in countries where the mortality rates were relatively high. (B) Daily number of cases and deaths in countries where the mortality rates were relatively low. Black dots represent observations, blue and orange curves represent the mean estimation results of daily cases and deaths; light orange and light blue shaded areas represent their 95% credible intervals. Daily mean estimation of cases, deaths and their credible intervals were obtained from the PMCMC posterior samples.

Generally, the estimated iCFRs in most of the countries decreased rapidly after reaching the maximum value in late March or early April and became more stable after then (Figure 3); however, variations still exist among countries. Among the four high mortality countries, iCFRs appeared to increase again slowly after reaching their minimum values (Figure 3A): the rates increased again from the minimum values 0.12 to 0.14 in the United Kingdom; 0.11 to 0.13 in Italy; 0.18 to 0.20 in France; and 0.09 to 0.12 in Spain. iCFRs among low mortality countries generally demonstrated a decreasing trend or maintained at low values (Figure 3B): the rates maintained between 0.05 and 0.06 in Germany; 0.07 and 0.08 in the Netherlands. In Sweden and Romania, after reaching their peaks in mid-April, the iCFR continued decreasing from 0.10 to 0.03 and from 0.07 to 0.03, respectively. In order to explore the association between weather conditions and the iCFR without the impacts of changes in NPIs, only the period when the value of Re remained below 1.5 was used. For most countries, Re reduced rapidly before April and fluctuated around 1 after then (Figure 3).

**Figure 3:**
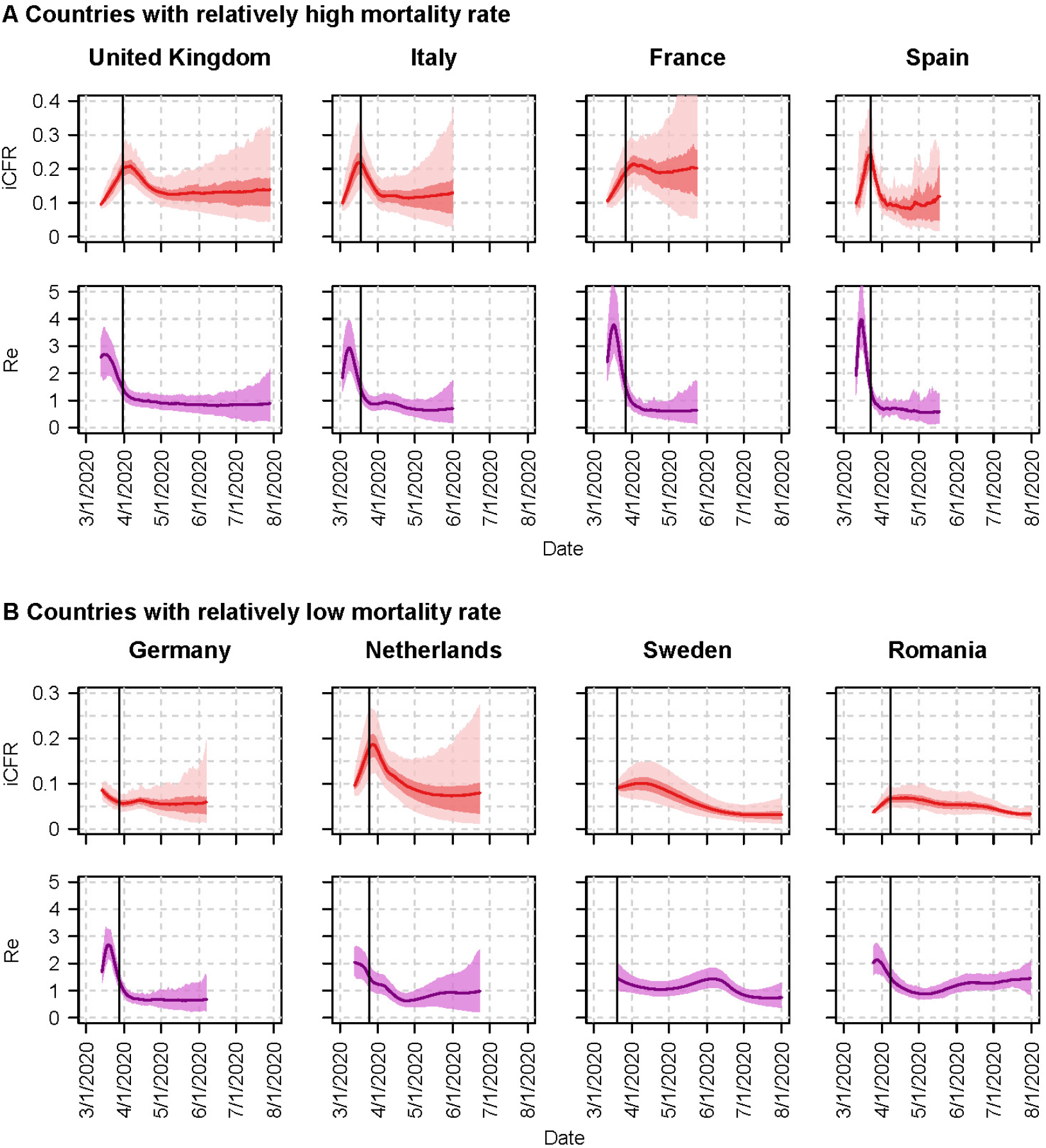
Model-estimated iCFR and Re in the eight European countries. (A) Daily iCFR and Re in countries where the mortality rates were relatively high. (B) Daily iCFR and Re in countries where the mortality rates were relatively low. Red and purple curves represent the estimated mean iCFR and Re, light red and dark red shaded areas represent the 95% and 50% credible intervals for iCFR. Purple shaded areas represent the 95% credible intervals for Re. The black vertical lines refer to the dates when Re reduced to below 1.5. iCFRs after these dates were used for estimating the effects of weather conditions. Daily mean iCFR, Re, and their credible intervals were obtained from the PMCMC posterior samples.

### 3.2. Weather effects on iCFRs

Daily mean temperatures among these European countries gradually increased during the first wave of the pandemic (Figure 4). Daily relative humidity in low mortality countries generally increased (Figure 4B), but the increase was not clearly observed in high mortality countries (Figure 4A). During the study period, the median daily temperatures ranged between 10.4°C and 16.5°C among the eight countries, and the median relative humidity ranged between 61.8% and 75.4% (Supplementary Table 3). Furthermore, most countries had temperatures ranging from 4°C to 20°C, and relative humidity ranged from 40% to 87%. The temperature changed the most in Sweden from -2°C to 21°C, and changed the least in Spain from 7°C to 20°C. Romania had the largest change in the relative humidity (i.e. 30% to 87%), and Spain had the smallest (i.e. 63% to 86%).

**Figure 4:**
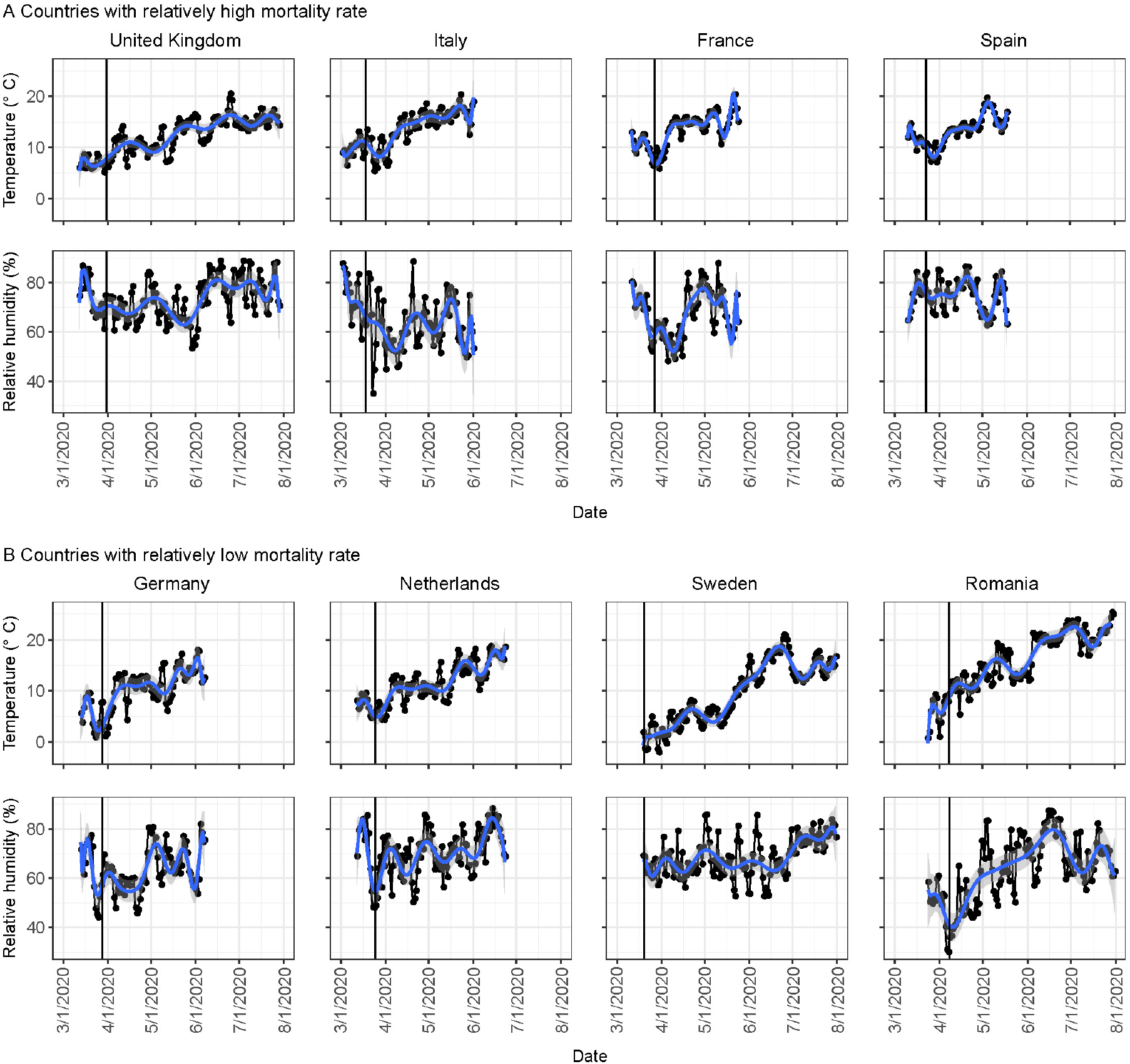
Daily mean temperature and relative humidity for the eight European countries. (A) Daily mean temperature and humidity in countries where the mortality rates were relatively high. (B) Daily mean temperature and humidity in countries where the mortality rates were relatively low. The blue curve represents the trend of daily temperature and humidity, which was obtained from a smoothing curve (a 18th order polynomial function). The grey shadows represent 95% credible intervals for the trends. The black vertical lines refer to the dates when Re was reduced to below 1.5.

The DLNM model with both temperature and relative humidity as predictors was selected as the best-fitting model for assessing the effects of environmental conditions (see Methods, Supplementary Table 4 and Supplementary Table 5). Figure 5B depicted the associations between temperatures and risk of death with a median temperature of 11°C as the reference. Lower temperatures, especially when temperatures were below 8°C, were more likely to increase COVID-19 iCFRs. We found that the estimated odds ratio (OR) of fatality peaked at virus exposure time when the temperature was low (OR=1.29 (95% CI: 1.23, 1.35) at -0.1°C). However, surprisingly the OR reached a second peak value one day after symptom onset with similar temperature (OR=1.12 (95% CI: 1.08, 1.16) at 0.1°C, see Figure 5B). The lowest OR occurred at two days after symptom onset (OR=0.71 (95% CI: 0.63, 0.80) at 21.7°C), and the second-lowest was observed at virus exposure time (OR=0.81 (95% CI: 0.71, 0.92) at 23.2°C).

**Figure 5:**
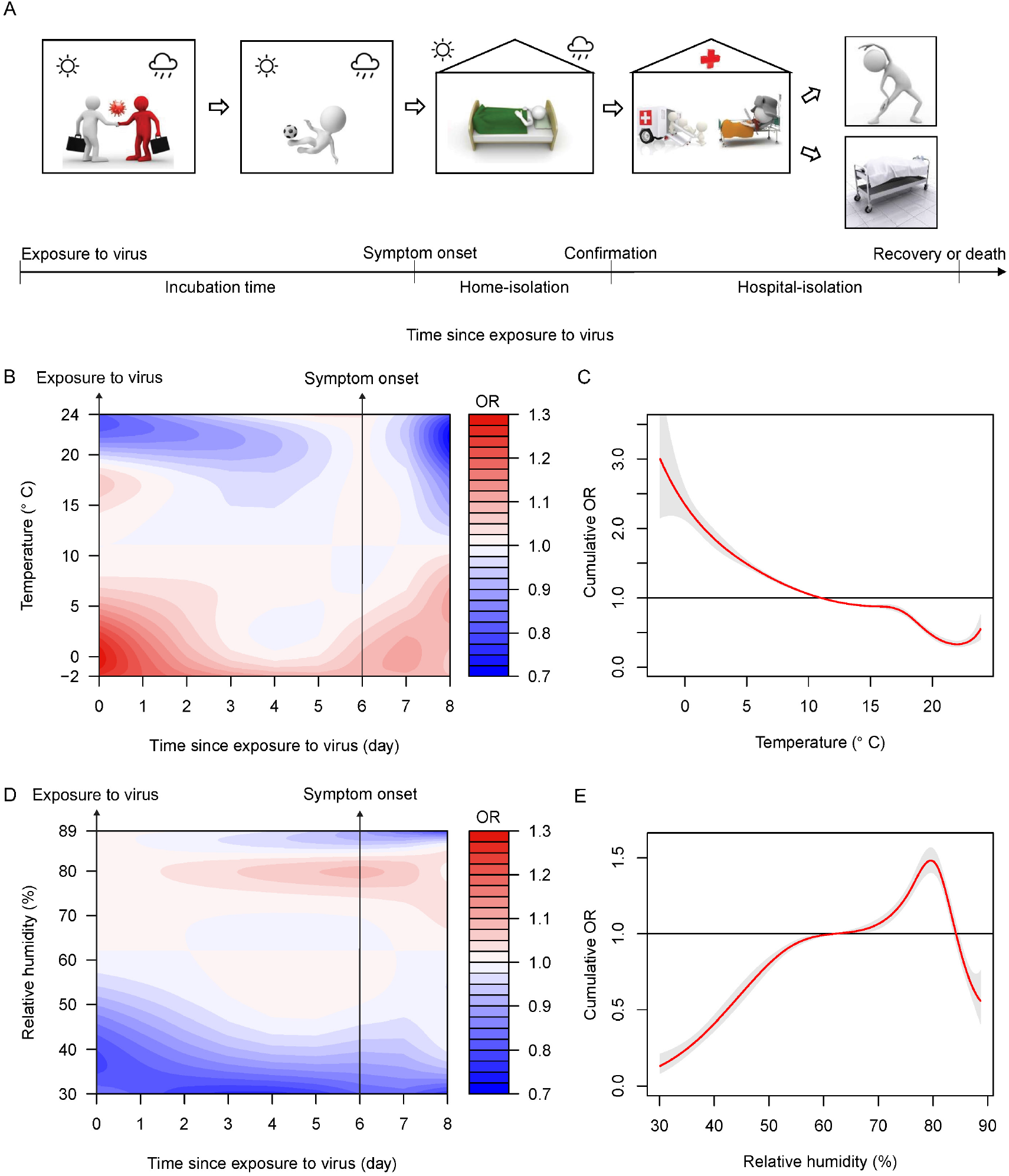
The effect of weather conditions on the COVID-19 fatality rate. (A) The timeline of COVID-19 infection course while taking account of the effects of weather conditions (i.e. temperature and relative humidity). The duration of home-isolation was approximately equal to the confirmation delay because many cases were isolated at hospitals after being confirmed. The duration of hospital-isolation for cases who later died was indicated by the infection outcome delay. Weather conditions mainly affected infected individuals during the early infection period including exposure to virus, incubation time and home-isolation period. During hospital-isolation period, indoor temperature and humidity were controlled by hospitals. (B and D) Relationships between weather conditions and the OR of fatality at different time point between exposure to virus and two days after symptom onset. Redder colours indicate higher OR. 11°C of the temperature and 62% of the relative humidity were used as references. (C and E) The estimated cumulative effects of the temperature and the relative humidity on the fatality. The red lines are the mean ORs, and the grey shaded areas are the 95%credible intervals.

These results suggest that both the initial viral load during the virus exposure time, and the immune responses at approximately a few days after symptom onset were affected by environmental temperatures. Furthermore, results showed the impacts of temperatures on the risk of death were greater at virus exposure time and few days after symptom onset than other periods during early infection stages. For example, a decrease from 5°C to 0°C at one day after symptom onset increased the risk of deaths significantly (OR increased from 1.03 to 1.07; see Figure 5B). This increase was significantly greater than during the presymptomatic transmission period (e.g. 6-folds greater than that) at three days before symptom onset (OR only increased from 1.006 to 1.012).

The overall cumulative OR of temperature during the early infection stages was calculated by summing the effect of each time point between exposure to the virus and two days after symptom onset. A negative relationship between temperature and the OR of fatality was observed over the range of -2°C to 22°C (Figure 5C). With exposure to a warm temperature of 24°C, the cumulative ORs during the first two days after symptom onset was 0.79. In order to check whether the results are caused by autocorrelation in weather, we further performed Fourier analysis. We found that temperature in Italy has a more distinct pattern of 7 or 8-day periodicity. There is no similar pattern in temperature among other countries or in relative humidity (Supplementary Figure 2).

Figure 5D showed the associations between the humidity and risk of death, with a relative humidity of 62% as the reference. The highest OR (1.08, 95% CI: 1.07, 1.10) was observed at 79.6% relative humidity at symptom onset time. Figure 5E showed the cumulative OR increased when the humidity raised from 30% to 80%. However, the cumulative OR clearly reduced when the humidity increased from 80% to 89%. This reduction was mainly resulted from the effect of humidity after symptom onset (see Figure 5D). The cumulative ORs for the first two days after symptom onset was 0.66, with the exposure of relative humidity of 89%.

We checked the robustness of the estimated associations between weather conditions and the risks of death after re-fitting the model to data during different periods of time (i.e. from March to April, March to May, and March to June). In addition, to verify if the selected model is affected by the sample size, we re-fitted the model using smaller-size data sets (i.e. 5%, 10%, 15% and 20% of samples were removed from the full dataset). In both cases, we found that the effects of weather conditions on the risk of death were generally consistent (see Supplementary Figure 3 and Supplementary Figure 4).

In summary, the variations of iCFRs within the eight European countries were associated with changes in weather conditions. Furthermore, the OR of fatality was clearly associated with the temperature and humidity that patients were exposed to at two distinct infection stages: virus exposure and after symptom onset.

## 4. Discussion

Although previous studies have demonstrated the impact of weather conditions (e.g. temperature and humidity) on COVID-19 deaths [29, 30, 13], it is still unknown how such conditions affect COVID-19 fatality risk during the infection progress. This was the first study focused on the impact of weather exposures at the early infection course on the probability of death. We found that the temperature and humidity affected the risk of death significantly not only at virus exposure time but also after symptom onset (Figure 5), which suggests that environmental conditions may influence both the initial viral load and an individual’s immune response to the virus (presumably through the innate immune system). These findings were obtained from distributed lag nonlinear models [27] with the iCFR estimated using a stochastic disease transmission model that accounted for delays in infection confirmation and infection outcome.

During the first epidemic wave in Europe, certain countries suffered high mortality rates. We found that warm conditions reduced the risk of deaths especially when the temperature was greater than 15°C. Among the study countries, Romania showed a temperature warmer than this threshold for a long period of time (more than half of the study period). Sweden and Netherlands also showed a warmer temperature for several days. In addition, our results showed that the risk of death was low when the relative humidity ranged below 50% (Figure 5E). Germany and Romania had humidity below 50% for a longer time, which may be a possible reason to explain why they experienced lower iCFR. A negative relationship between the relative humidity and iCFR was observed when the humidity was larger than 80%. Our results may help explain diverging patterns in previous studies, namely that humidity and case fatality are negatively correlated in humid areas [29], but positively correlated in arid regions [30].

How extremely high humidity (> 80% in Figure 5E) can affect the severity of COVID-19 remains largely unknown. Humidified air has been previously found to reduce the severity of respiratory infection of influenza in mice, through increasing the function of mucosal barrier [31]. Similarly, a recent study proposed that the use of face masks is linked to the reduced severity of COVID-19 infections because the humidity of inspired air increases [5]. The beneficial effect of extremely high humidity (>80%) that was observed is likely due to a similar mechanism. Mucus layers constitute a biochemical barrier to inhibit pathogen penetration [32]. A well-hydrated mucus layer ensures the continuous flow of mucus, responsible for removing pathogens from the airways and lungs [33]. Although the association between weather and COVID-19 related deaths has been previously reported [29, 30, 13], questions such as when (e.g. which infectious stage) and how these factors affect disease fatality have not been clarified. Many infected individuals, after being contacttraced or developing symptoms, are isolated at home or quarantine/isolation centers. During this period, certain environmental risk exposures can be manually controlled or avoided through an air-conditioner or other devices. We found that exposure to warmer temperatures (24°C) during the first two days after symptom onset can roughly reduce the iCFR by 19% compared to exposure to the reference temperature (11°C). Consistent with recent laboratory experiments showing that wearing face masks can increase the humidity of inspired air and reduce the severity of COVID-19 [5], we found that exposure to high relative humidity (89%) during the first few days after symptom onset can reduce the CFR by 31% compared to the reference level (62% relative humidity). Based on all these results, therefore, we recommend adopting certain individual preventive measures: 1. to stay in a proper warm place after symptom onset; 2. to wear a face mask to increase the humidity of inspired air. However, further epidemiological observational studies (e.g. case-control study) in different population and environments are still needed to determine optimal indoor environmental conditions after symptoms appeared.

In addition to the evidence that low temperatures increased the stability and viability of the virus, inhalation of cold air at the initial virus exposure time, can make the upper airway more suitable for viral replication [34, 4], resulting in large viral load, and potentially more severe adverse outcomes. On the other hand, how the temperature that patients were exposed to after symptom onset affect immune responses is still unclear. This can be explained by some hypotheses of innate immunity. Macrophages, which produces cytokines and chemokines, have been found to increase in the lower airways after exposure to cold air [35]. IL-6 and TNF-alpha, the cytokines that play important roles in mediating SARS-CoV-2-associated cytokine storms [36, 37] have been reported to increase after cold exposure [38]. The activation of these factors usually begins within a week after symptom onset [3]. In general vagus nerve circuits can regulate cytokines release in macrophages to prevent potentially damaging inflammation [39, 40, 8]. However, exposure to cold stimulates cold receptors of the skin, which increase sympathetic tone and might antagonistically reduce this vagus activity [7, 41]. The overall net effect might reduce the neural mediated anti-inflammatory activity.

Over reaction of innate immunity may cause immune system dysregulation and severe adverse outcomes (i.e. acute respiratory distress syndrome, systemic inflammatory response syndrome, and cardiac failure [42]). These hypotheses of innate immunity suggest that there exists a link between temperature exposure in the early infection stages and COVID-19 disease severity.

Some limitations exist in this study. First, the effects of other interventions (e.g. increasing number of PCR tests performed and improvement of medical treatment) on the iCFR were not considered. There are minor variations in the weekly number of COVID-19 tests conducted in the eight European countries during the study period [43]. To avoid the effects caused by the variation of medical treatment and NPIs, we used the data during the first wave of the pandemic when the changes in interventions were minor (when Re was stable and near one). Second, the data of indoor temperature and humidity are not available. It is reasonable to believe that the house indoor temperature and humidity are largely affected by the weather in Europe where air-conditioners or heaters are not frequently used at home during our study period. Thus, there is a monotonic relationship between house indoor condition and weather. Third, we cannot rule out the effect of certain confounding factors related to personal behaviours. (e.g. the dry air in winter can irritate people’s airways, which triggers more people to be cautious and reduce outdoor activities). In addition, because in some countries, many cases were confirmed and isolated in hospitals around two days after symptom onset on average (see Figure 1B), hence their environmental exposure would mainly be determined by the hospital air conditioning system. Therefore, we only assess the impact of weather exposure no longer than that time.

## Data Availability

Daily reported COVID-19 cases and deaths in these eight European countries from 16th February to 31st June 2020 were collected from the Our World in Data (https://github.com/owid/covid-19-data/tree/master/public/data). Daily weather data for the eight European countries were collected from the European Climate Assessment and Dataset (https://www.ecad.eu/) and United States National Oceanic and Atmospheric Administration (https://www.ncei.noaa.gov/access/search/data-search/global-summary-of-the-day).

## Ethics approval

Ethics approval is not needed as the study uses publicly available country-level (aggregated) morbidity, mortality, and weather data.

## Data availability

Daily reported COVID-19 cases and deaths in these eight European countries from 16th February to 31st June 2020 were collected from ‘Our World in Data’ (https://github.com/owid/covid-19-data/tree/master/public/data). Daily weather data for the eight European countries were collected from the European Climate Assessment and Dataset (https://www.ecad.eu/) and United States National Oceanic and Atmospheric Administration (https://www.ncei.noaa.gov/access/search/data-search/global-summary-of-the-day). The data and materials are available from the corresponding author on reasonable request.

## Acknowledgements

We thank Dr. Kwan Ting Chow at City University of Hong Kong and Dr. Liao Ben-Yang at National Health Research Institutes for their valuable comments. We thank Dr. Yun Huang at Sun Yat-sen University, Dr. Lindsey Wu at London School of Hygiene & Tropical Medicine, Dr. Rahul Subramanian at University of Chicago, M. Pear Hossain, Zhaojun Ding, and Dr. Sir Colin Blakemore at City University of Hong Kong for their suggestions and contributions in the preparation of the manuscript. We thank Dr. Antonio Gasparrini at London School of Hygiene & Tropical Medicine for his suggestions on the verification of sample size in the DLNM model.

## Author contributions

HY and JL designed the study. JL collected, analyzed, and modelled the data. JL and HY wrote the paper.

## Supplementary Materials

## 1. Supplementary Methods

### 1.1. Data

For countries (the United Kingdom, France and Romania) lacking relative humidity data, we calculated relative humidity values using the ratio of the actual water vapor pressure (*P*_*w*_) divided by the saturation water vapor pressure (*P*_*ws*_) using the following formula [1]:

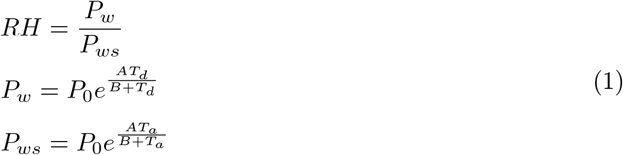

Where *RH* denoted the relative humidity. *P*_0_ denoted the saturation vapor pressure as 6.11 MB with a reference temperature of 273.15°K. A (17.43) and B (240.73) are constant values [1]. *T*_*d*_ is the dew point collected from the United States National Oceanic and Atmospheric Administration [2], and *T*_*a*_ is the observed temperature. In this study, the daily mean temperatures and relative humidity values for each country were calculated as the average records from all monitoring stations.

### 1.2. SEIR stochastic model

There were several assumptions for the SEIR model: (1) susceptible individuals became exposed after contacting with the virus; (2) exposed individuals could be divided into two groups, non-infectious exposed individuals and infectious pre-symptomatic individuals [3]; and (3) all hospital confirmed cases were isolated and could not transmit the disease. Based on these assumptions, our SEIR model was derived as follows:

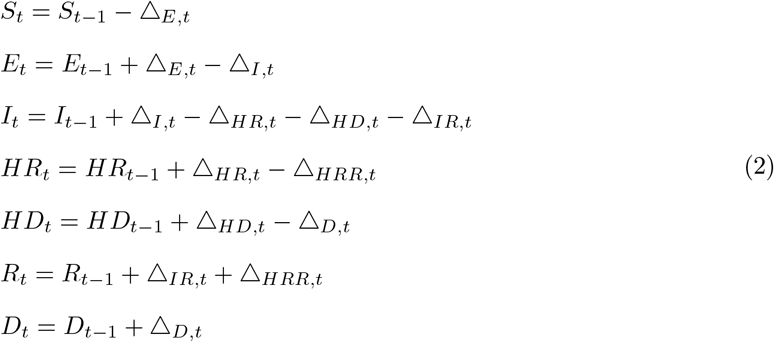

Where Δ_*E,t*_ represented the number of newly exposed individuals during the time interval t-1 to t, Δ_*I,t*_ was the number of newly symptom-onset infections, Δ_*HR,t*_ was the number of newly hospital-confirmed cases who later recovered, Δ_*HD,t*_ was the number of newly hospital-confirmed cases who later died, Δ_*IR,t*_ was the number of newly recovered cases who were not reported (confirmed) by hospitals, Δ_*HRR,t*_ was the number of newly recovered cases that were reported (confirmed) by hospitals, and Δ_*D,t*_ was the number of newly reported deaths. We assumed that Δ_*E,t*_, Δ_*I,t*_,Δ_*HR,t*_ + Δ_*HD,t*_, Δ_*IR,t*_, Δ_*HRR,t*_, and Δ_*D,t*_ followed Poisson distributions, and Δ_*HD,t*_ followed a binomial distribution with the rate parameter as the *iCFR*_*t*_:

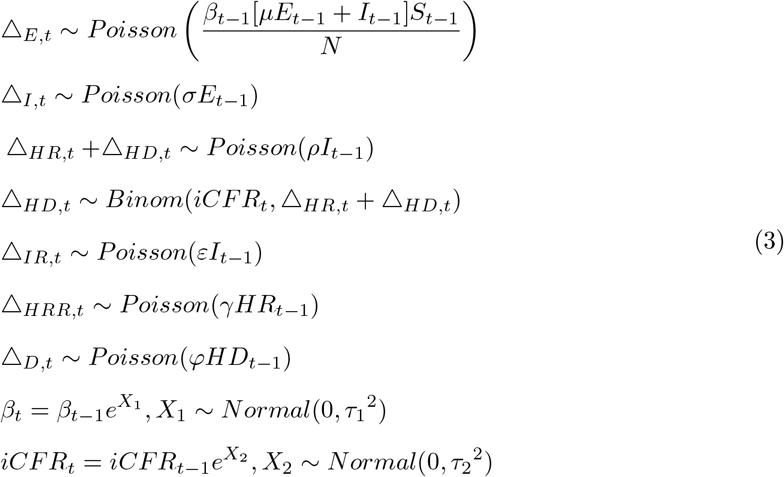

where *β*_*t*_ is the transmission rate; *μ* is the proportion of infectious pre-symptomatic individuals among all exposed individuals, calculated as 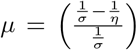 [3], in which *η* was the rate at which exposed individuals become infectious pre-symptomatic infections (1*/η* was the latent period), and *σ* was the rate at which exposed individuals become symptom-onset infections (1*/σ* was the incubation period); *ρ* was the case confirmation rate (1*/ρ* was the time between symptom onset and case confirmation), *iCFR*_*t*_ was the iCFR at time t, *γ* was the recovery rate of hospital-confirmed cases (1*/γ* was the time between case confirmation and recovery), *ε* was the recovery rate of cases who were not reported (confirmed) by hospitals, and *φ* was the death rate (1*/φ* was the time between case confirmation and death). During model fitting, we assumed that *β*_*t*_ and *iCFR*_*t*_ followed Brownian motion by time [4] with the standard deviation parameters as *τ*_1_ and *τ*_2_. *τ*_1_ was set as 0.25 for Sweden’s, Romania’s and the United Kingdom’s model; as 0.3 for Germany’s and France’s model; as 0.35 for Netherlands’ and Italy’s model; as 0.4 for Spain’s model. *τ*_2_ was set as 0.25 for Sweden’s and Romania’s model; as 0.3 for the United Kingdom’s and Germany’s model; as 0.35 for Italy’s, France’s and Netherlands’ model; as 0.4 for Spain’s model. The values of *τ*_1_ and *τ*_2_ were chosen by the smallest Watanabe–Akaike Information Criterion (WAIC) [5, 6].

To map our model outputs to actual observed data, we constructed an observation model (equation (4)), assuming the variations between the actual data and our model predictions follow Gaussian distributions. As equation (4) showed, the likelihood of observing daily reported new cases (*Cases*_*rep,t*_) was calculated through a Gaussian distribution with the mean defined as the number of model predicted new-confirmed cases (Δ_*HR,t*_ + Δ_*HD,t*_) where *τ*_*c*_ (20% of the maximum of *Cases*_*rep,t*_) was the standard deviation. Similarly, the likelihood of observing daily new deaths (*Deaths*_*rep,t*_) was also calculated through a Gaussain distribution with the mean defined as the number of model predicted new deaths (Δ_*D,t*_), where *τ*_*d*_ (20% of the maximum of *Deaths*_*rep,t*_) was the standard deviation.

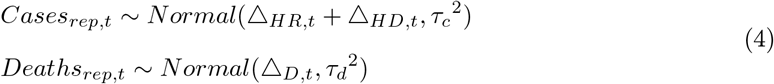

Based on the next-generation matrix (NGM) method [7], time-varied country-specific Re was calculated as the first eigenvector of the matrix *NGM*_*t*_ [8]:

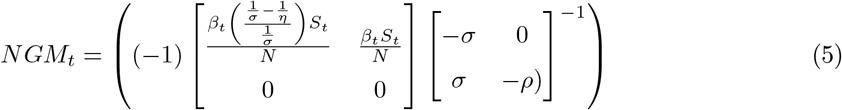

Prior distributions for the parameters were set as follows: for the latent period, 1*/η* ∼ U[2, 3] [9]; for the incubation period, 1*/σ* ∼ N[5.5, 0.25] [10]; for the time between symptom onset and case confirmation, 1*/ρ* ∼ U[2, 6] [11, 12]; for the recovery time of hospital-confirmed cases, 1*/γ* ∼ N[14, 0.01] [13]; for the recovery time of hidden cases who were not reported (confirmed) by hospitals, 1*/ε* ∼ U[16, 0.01] [13]; for the time between case confirmation and death, 1*/φ* ∼ U[7, 14] [14, 12].

### 1.3. DLNM model

To explore the effects of weather conditions during the early infection period, we defined the minimum lag times (*lag* = 0) as the second day after symptom onset, given some infected cases would be isolated in the hospitals two days after symptom onset (Supplementary Table 2) and the outdoor environment would not influence their infection process during that time; we set the maximum lag time as nine days (the length between exposure to the virus and two days after symptom onset, Supplementary Table 2). In this study, we used the odds ratio (OR) of fatality to explore the association of death and different levels of temperature/humidity. The OR of fatality was calculated as the ratio of the odds of fatality when subjects were exposed to a particular level of temperature/humidity to the odds of fatality when subjects were exposed to the reference temperature/humidity. Here, we choose a median reference temperature of 11°C and relative humidity of 62%.

The model was formulated as:

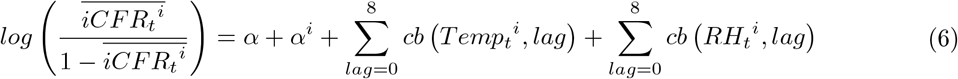

where 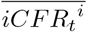 was the expected iCFR in country *i* at time *t*; *α* was the intercept; *α*^*i*^ represented country-specific random effects (e.g. local conditions in health care); *Temp*_*t*_^*i*^ was the daily mean temperature; *RH*_*t*_^*i*^ was the daily mean relative humidity value; *lag* was the lag time between exposure to the virus and two days after symptom onset; *cb (Temp*_*t*_^*i*^, *lag*) and *cb (RH*_*t*_^*i*^, *lag*) were cross basis functions [15], modeling weather exposure-response and lag-response relationships, with quadratic B-spline function knots placed at equal intervals in the weather range to permit sufficient flexibility in the tails. The Akaike information criterion (AIC) was used to test the model robustness using different degrees of freedom (knots). We found five degrees of freedom for temperature and relative humidity had the best performance in model fitting.

## 2. Country-specific effect on fatality rate

We used a generalized linear mixed model (i.e. supplementary Equation (6)) to incorporate the country-level variations as random effects (i.e. varying-intercept random model). To explore these random effects on fatality rates for different periods of time, we re-fitted the model using data during March to April, March to May, and March to June separately.

As shown in Supplementary Table 6, France had the largest intercept (i.e. country-specific effect on case fatality rate) when the full data set (i.e. March to July) was used. Italy had the second largest. Sweden had the lowest intercepts. According to the model formula in the supplementary Equation (6), while holding all other predictors (i.e. temperature and relative humidity) constant, the larger intercept indicates a higher risk of death.

After re-fitting data during different periods of time (i.e. from March to April, March to May, and March to June), we found that the effects of the country-level variations on fatality were similar. France consistently had the largest intercept (estimated mean intercept was -2.70 for March to April, -2.49 for March to May, -2.22 for March to June); Italy had the second-largest except for March to April (estimated mean intercept was -2.88 for March to May, -2.62 for March to June); and Germany and Sweden had smaller intercepts. Moreover, we checked the robustness of the estimated associations between weather conditions and the risks of death for these re-fitted models. As shown in Supplementary Figure 3, the relationships between temperature (and humidity) and risk of death risk were consistent when datasets between different time periods were used (i.e. March to April, March to May, March to June).

## 3. Verification of sample size in DLNM model

The sample size of each country of the full data set was shown in Supplementary Table 7. In order to verify the impact of sample size on the selected model (see equation (6) in supplementary methods), we validated the model by using different sample size: we randomly removed 5%, 10%, 15%, and 20% of the samples in each country from the full data set and re-fitted the model to the removed data set. We found that the effects of weather conditions on the risk of death were generally consistent even 20% of samples have been removed (Supplementary Figure 4). Warm temperature (>= 15°C) and low high humidity (<=50%) were able to reduce the risk of death compared to reference condition (i.e. temperature 11°C, relative humidity 62%). A negative relationship between the relative humidity and iCFR was observed at high humidity (>80%).

## Supplementary Figures

**Supplementary Figure 1:**
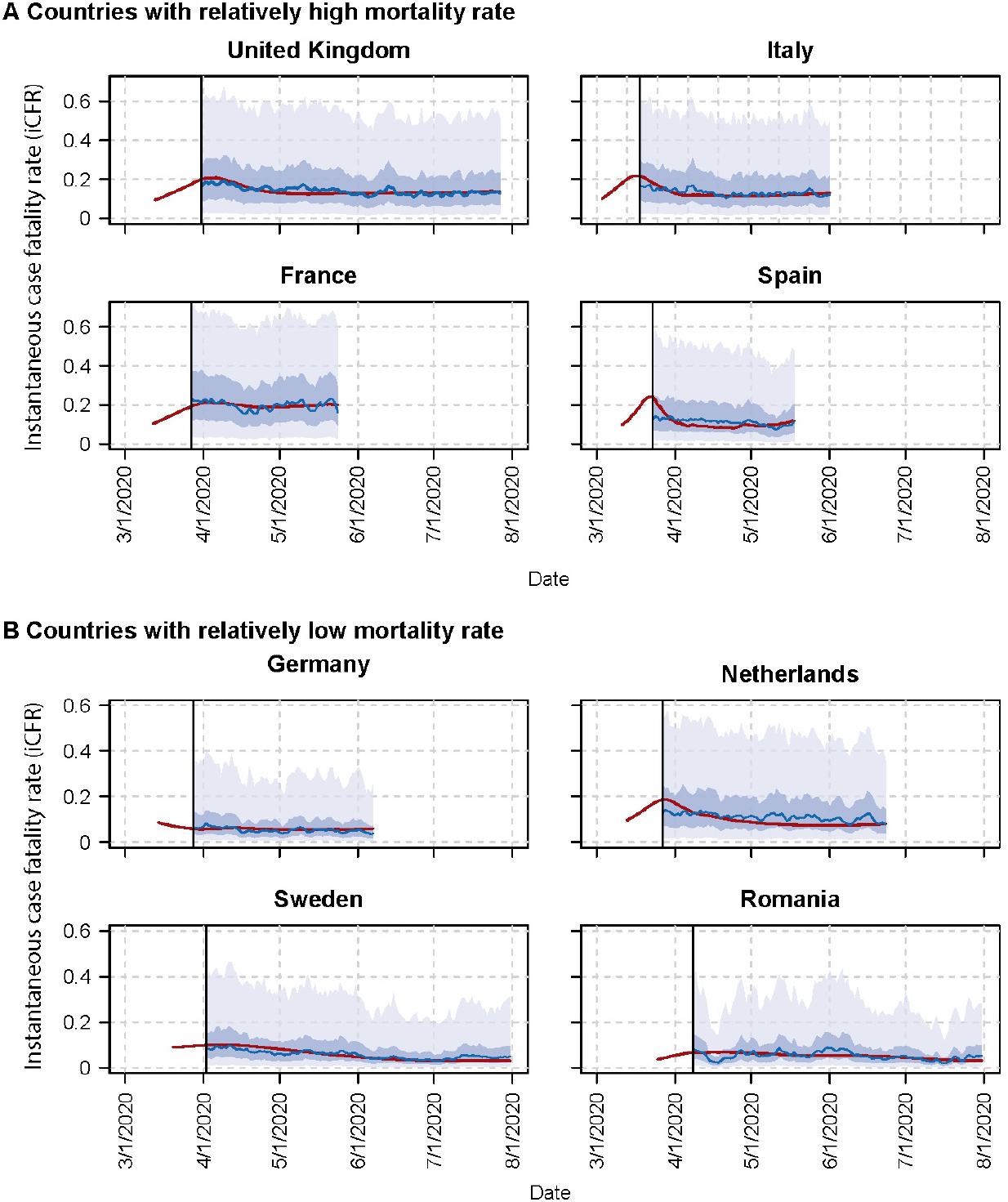
Prediction for the iCFR based on DLNM models in the eight European countries during the community outbreaks. The red curves represent the iCFR calculated by SEIR models. The blue curves represent the mean prediction of iCFR based on DLNM models. The light blue and dark blue shaded areas respectively represent the 95% and 50% prediction intervals.

**Supplementary Figure 2:**
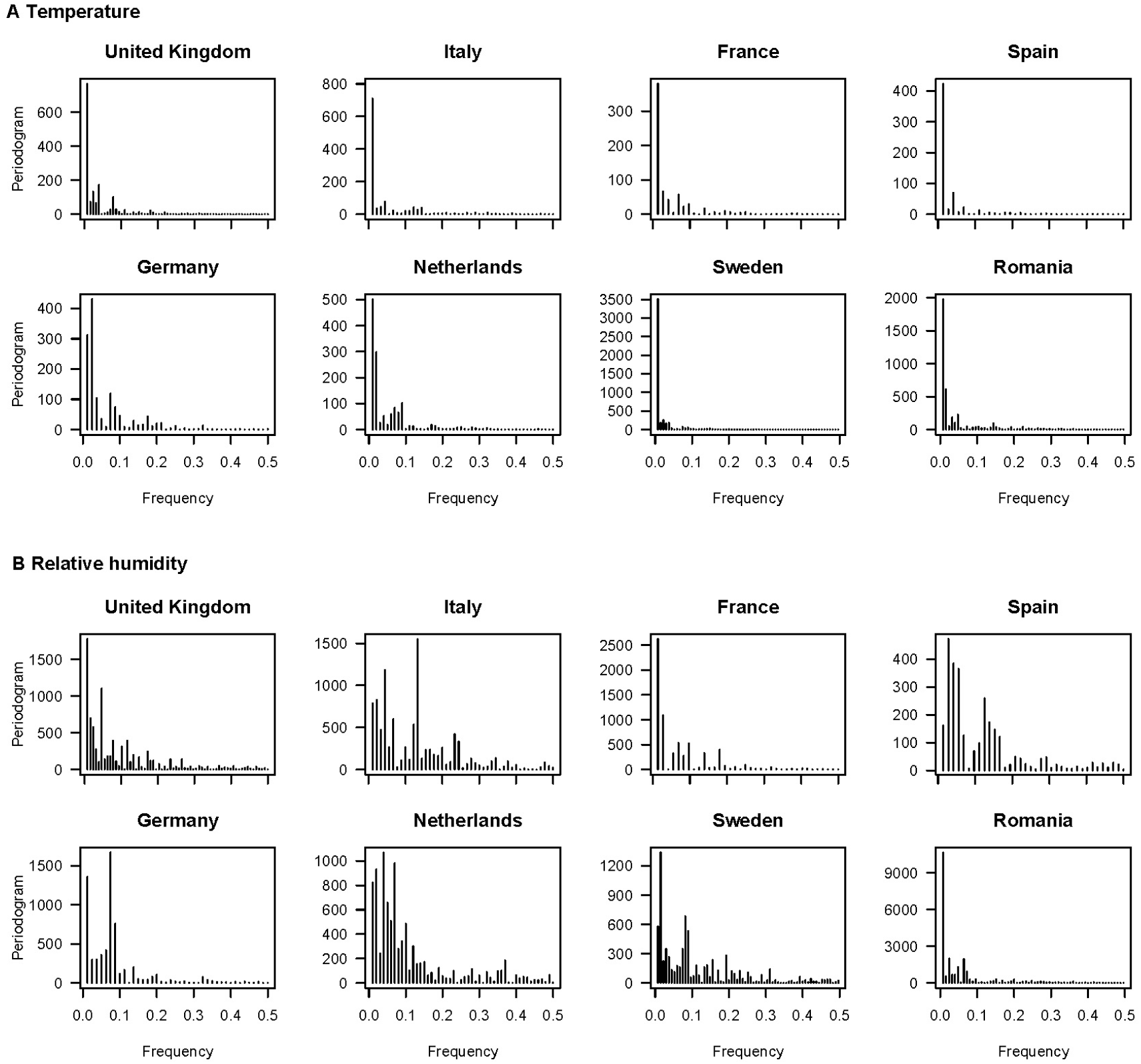
Periodogram plot of daily temperature and relative humidity among the eight countries.

**Supplementary Figure 3:**
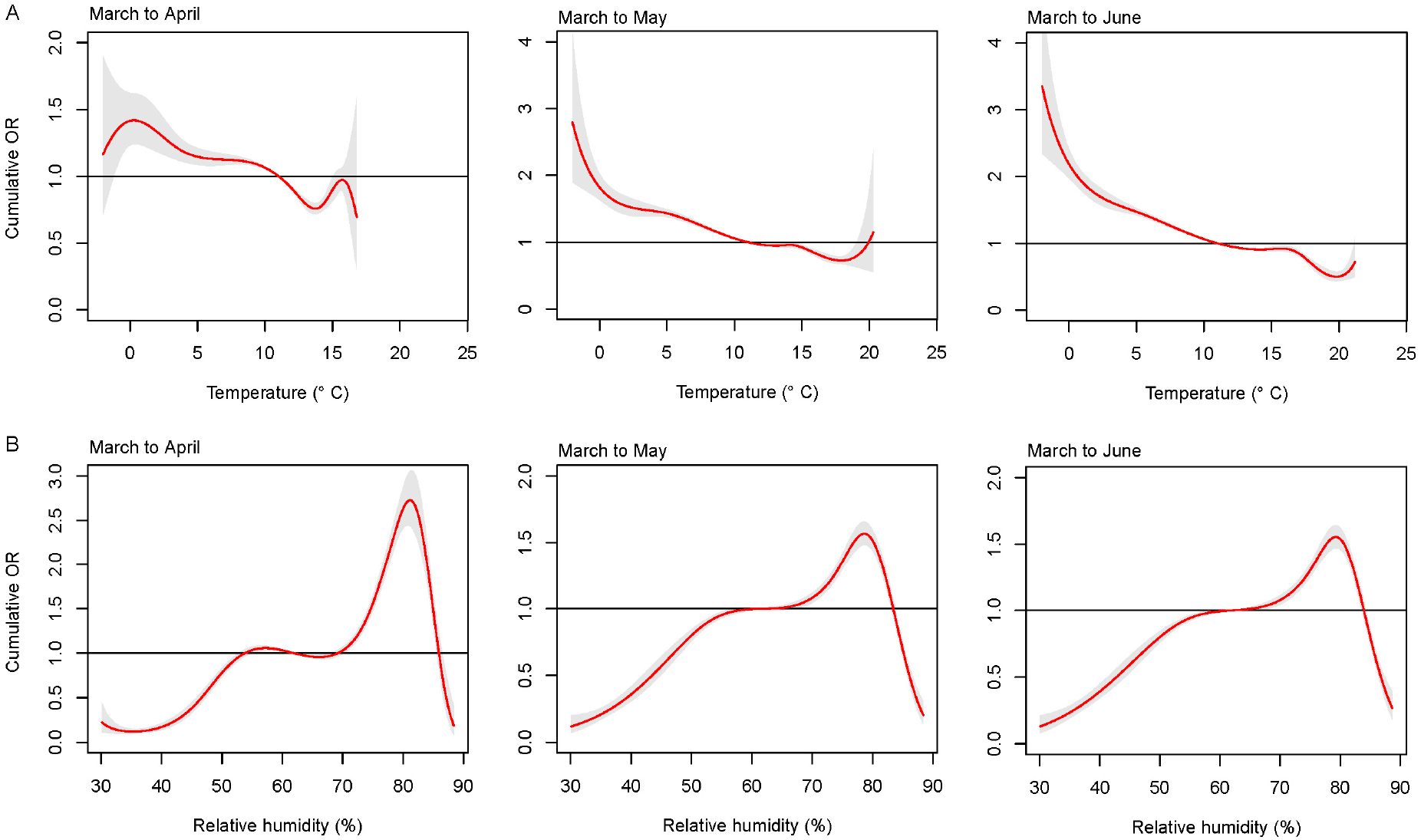
The estimated cumulative effects of the temperature and relative humidity on fatality rates. The red lines are the mean ORs, and the grey shaded areas are the 95% credible intervals.

**Supplementary Figure 4:**
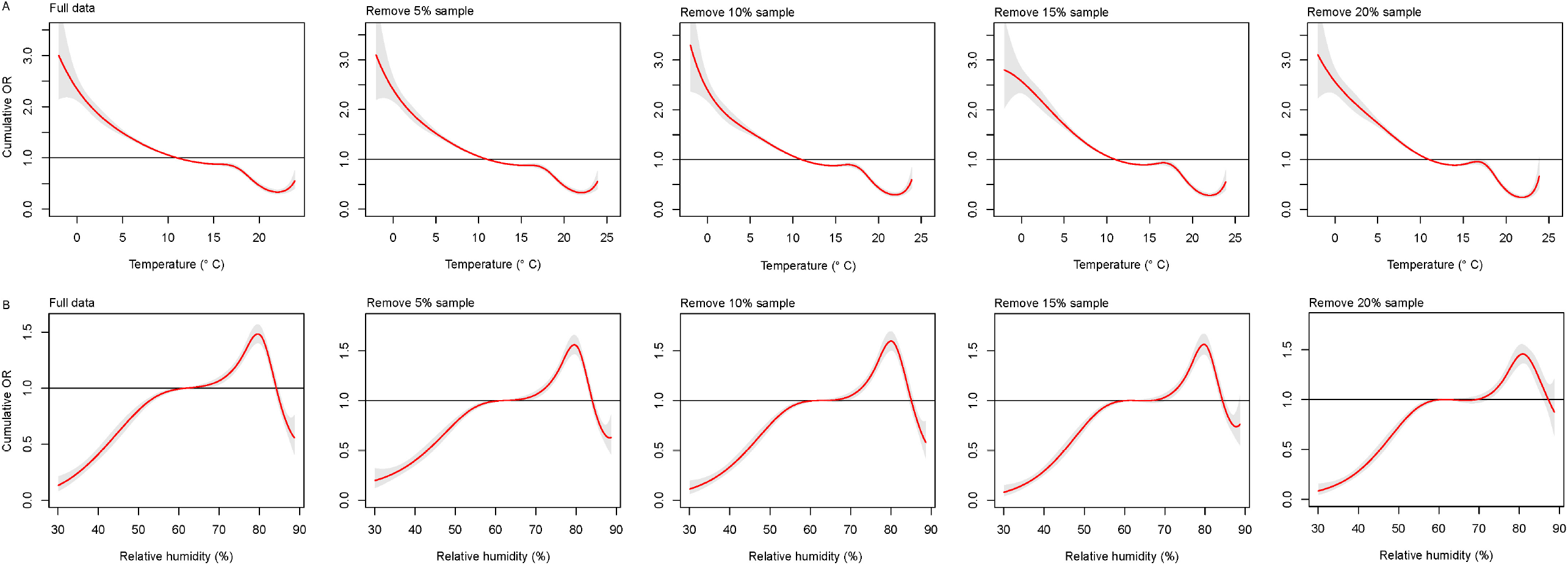
The estimated cumulative effects of the temperature and the relative humidity on fatality rates using different training data sets.

**Supplementary Table 1:** Parameter estimates for the SEIR epidemic model. The mean values and 95% credible intervals of the posterior distribution of each of the parameters are included. 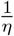 is the latent period, 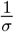 is the incubation period, 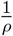 is the confirmation delay, 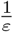 is the recovery time for un-reported cases (mainly asymptomatic cases), and 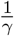 is the time from confirmation to recovery for hospital confirmed cases. 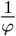 is the infection outcome delay. Gelman-Rubin convergence (GR) test was performed. Convergence occurs when the value of GR is close to 1.

| | $\frac{1}{\eta}$ | | $\frac{1}{\sigma}$ | | $\frac{1}{\rho}$ | | $\frac{1}{\varepsilon}$ | | $\frac{1}{\gamma}$ | | $\frac{1}{\varphi}$ | | WAIC |
| --- | --- | --- | --- | --- | --- | --- | --- | --- | --- | --- | --- | --- | --- |
|  | Mean<br>(95% CI) | GR | Mean<br>(95% CI) | GR | Mean<br>(95% CI) | GR | Mean<br>(95% CI) | GR | Mean<br>(95% CI) | GR | Mean<br>(95% CI) | GR |  |
| United Kingdom | 2.39<br>(2.02,2.97) | 1.01 | 5.71<br>(4.96,6.54) | 1.00 | 4.03<br>(2.42,5.62) | 1.01 | 16.00<br>(15.83,16.22) | 1.00 | 13.99<br>(13.78,14.14) | 1.06 | 7.92<br>(7.02,10.31) | 1.00 | 4256.8 |
| Italy | 2.28<br>(2.01,2.87) | 1.00 | 5.60<br>(4.66,6.48) | 1.01 | 3.00<br>(2.04,4.74) | 1.00 | 16.01<br>(15.83,16.21) | 1.00 | 13.98<br>(13.79,14.18) | 1.00 | 9.23<br>(7.09,12.98) | 1.01 | 2662.4 |
| France | 2.27<br>(2.00,2.81) | 1.01 | 5.28<br>(4.64,6.55) | 1.25 | 2.82<br>(2.01,4.82) | 1.00 | 15.96<br>(15.78,16.13) | 1.00 | 13.98<br>(13.82,14.25) | 1.03 | 9.00<br>(7.11,12.23) | 1.04 | 2247.2 |
| Spain | 2.13<br>(2.01,2.46) | 2.44 | 5.35<br>(5.22,5.47) | 1.11 | 2.44<br>(2.02,3.30) | 1.09 | 15.98<br>(15.85,16.09) | 1.00 | 14.04<br>(13.86,14.13) | 2.98 | 12.43<br>(10.51,13.59) | 1.13 | 2080.4 |
| Germany | 2.15<br>(2.00,2.60) | 1.14 | 5.47<br>(4.68,6.22) | 1.04 | 2.45<br>(2.01,3.57) | 1.00 | 15.98<br>(15.77,16.20) | 1.10 | 14.01<br>(13.86,14.17) | 1.21 | 12.10<br>(8.92,13.93) | 1.00 | 2364.6 |
| Netherlands | 2.56<br>(2.03,2.98) | 1.00 | 5.84<br>(4.95,6.67) | 1.01 | 5.42<br>(4.18,5.98) | 1.00 | 16.00<br>(15.79,16.18) | 1.00 | 13.99<br>(13.81,14.19) | 1.00 | 9.84<br>(7.20,13.62) | 1.00 | 2390.8 |
| Sweden | 2.60<br>(2.06,2.98) | 1.00 | 5.17<br>(4.23,6.12) | 1.01 | 3.72<br>(2.09,5.79) | 1.00 | 16.01<br>(15.82,16.21) | 1.00 | 14.00<br>(13.79,14.21) | 1.00 | 10.33<br>(7.17,13.78) | 1.01 | 3648.5 |
| Romania | 2.29<br>(2.01,2.86) | 1.00 | 5.09<br>(4.23,5.89) | 1.06 | 2.89<br>(2.02,4.63) | 1.00 | 16.01<br>(15.83,16.21) | 1.00 | 13.99<br>(13.79,14.21) | 1.00 | 8.12<br>(7.02,10.83) | 1.00 | 2718.5 |

## Supplementary Talbes

**Supplementary Table 2:** Summary statistics of daily number of cases, and deaths per 1 million people in the eight European countries.

|  | Minimum | 25% | Median | 75% | Maximum |
| --- | --- | --- | --- | --- | --- |
| **Cases** |  |  |  |  |  |
| United Kingdom | 1 | 11 | 21 | 53 | 82 |
| Italy | 5 | 14 | 38 | 66 | 109 |
| France | 2 | 9 | 23 | 40 | 113 |
| Spain | 5 | 25 | 63 | 105 | 196 |
| Germany | 3 | 8 | 14 | 41 | 76 |
| Netherlands | 4 | 10 | 16 | 47 | 77 |
| Sweden | 10 | 38 | 55 | 73 | 282 |
| Romania | 6 | 10 | 16 | 21 | 70 |
| **Deaths** |  |  |  |  |  |
| United Kingdom | 0 | 1 | 3 | 8 | 18 |
| Italy | 0 | 3 | 6 | 9 | 16 |
| France | 0 | 2 | 4 | 8 | 30 |
| Spain | 0 | 4 | 8 | 13 | 20 |
| Germany | 0 | 0 | 1 | 2 | 4 |
| Netherlands | 0 | 1 | 2 | 6 | 14 |
| Sweden | 0 | 2 | 4 | 8 | 18 |
| Romania | 0 | 1 | 1 | 1 | 2 |

**Supplementary Table 3:** Summary statistics of daily weather conditions in the eight European countries.

|  | Minimum | 25% | Median | 75% | Maximum | Mean±SD |
| --- | --- | --- | --- | --- | --- | --- |
| Temperature (°C) |  |  |  |  |  |  |
| United Kingdom | 5.14 | 9.21 | 13.08 | 15.04 | 20.51 | 12.15±3.59 |
| Italy | 5.33 | 10.78 | 13.93 | 16.4 | 20.34 | 13.36±3.63 |
| France | 5.85 | 10.72 | 13.69 | 15.55 | 20.37 | 13.24±3.29 |
| Spain | 7.13 | 11.35 | 13.44 | 14.6 | 19.73 | 13.19±2.84 |
| Germany | 0.90 | 7.79 | 10.77 | 12.76 | 17.99 | 10.13±4.12 |
| Netherlands | 4.33 | 8.2 | 11.28 | 14.19 | 18.72 | 11.39±3.91 |
| Sweden | -2.07 | 4.81 | 10.38 | 14.52 | 20.99 | 9.56±5.95 |
| Romania | 0.72 | 11.84 | 16.51 | 20.63 | 25.37 | 15.73±5.74 |
| Relative humidity(%) |  |  |  |  |  |  |
| United Kingdom | 53.29 | 66.78 | 73.09 | 79.33 | 88.78 | 73.28±8.17 |
| Italy | 34.89 | 55.66 | 63.46 | 72.04 | 88.42 | 64.12±10.92 |
| France | 48.11 | 59.99 | 66.84 | 74.56 | 87.8 | 66.92±9.56 |
| Spain | 62.63 | 68.56 | 75.35 | 79.56 | 86.38 | 74.66±6.47 |
| Germany | 44.04 | 57.69 | 61.84 | 71.51 | 81.79 | 63.46±9.25 |
| Netherlands | 48 | 64.71 | 70.83 | 77.66 | 88.22 | 70.39±9.48 |
| Sweden | 52.62 | 62.21 | 67.35 | 74.59 | 85.68 | 68.36±7.9 |
| Romania | 30.03 | 53.46 | 63.3 | 73.02 | 87.31 | 63.4±13.18 |

**Supplementary Table 4:** DLNM Model comparison based on different weather predictors.

| Model | AIC | BIC | Loglik |
| --- | --- | --- | --- |
| $\log(\frac{iCFR_t^i}{1-iCFR_t^i}) = \alpha + \alpha^i$ | 12857.2 | 12866.4 | -6426.6 |
| $\log(\frac{iCFR_t^i}{1-iCFR_t^i}) = \alpha + \alpha^i + \sum cb(Temp_t^i)$ | 10519.2 | 10637.8 | -5233.6 |
| $\log(\frac{iCFR_t^i}{1-iCFR_t^i}) = \alpha + \alpha^i + \sum cb(RH_t^i)$ | 11921.4 | 12040.0 | -5934.7 |
| $\log(\frac{iCFR_t^i}{1-iCFR_t^i}) = \alpha + \alpha^i + \sum cb(Temp_t^i) + \sum cb(RH_t^i)$ | 9738.3 | 9966.4 | -4819.1 |

**Supplementary Table 5:** The estimation of the fixed intercept and coefficients in DLNM coupled with a mixed-effect model. The variables of cb.temperaturev1.l1 to cb.humidityv6.l4 are generated by the cross basis functions in DLNM.

| Variable | Estimate | Std. Error | z value | P-value |
| --- | --- | --- | --- | --- |
| Intercept | -3.1699289 | 0.355785 | -8.91 | <0.001 |
| cb.temperaturev1.l1 | 0.1411383 | 0.1275635 | 1.106 | 0.268547 |
| cb.temperaturev1.l2 | -0.3438178 | 0.1099579 | -3.127 | 0.001767 |
| cb.temperaturev1.l3 | -0.0825041 | 0.1148768 | -0.718 | 0.472636 |
| cb.temperaturev1.l4 | 0.1213828 | 0.1081841 | 1.122 | 0.261862 |
| cb.temperaturev2.l1 | -0.2599674 | 0.0972192 | -2.674 | 0.007494 |
| cb.temperaturev2.l2 | 0.0471271 | 0.0769912 | 0.612 | 0.540465 |
| cb.temperaturev2.l3 | -0.0306237 | 0.0892278 | -0.343 | 0.731442 |
| cb.temperaturev2.l4 | -0.3038603 | 0.0877978 | -3.461 | <0.001 |
| cb.temperaturev3.l1 | -0.0106087 | 0.1064286 | -0.1 | 0.920599 |
| cb.temperaturev3.l2 | -0.0580889 | 0.0902724 | -0.643 | 0.519909 |
| cb.temperaturev3.l3 | -0.3593119 | 0.0966693 | -3.717 | <0.001 |
| cb.temperaturev3.l4 | -0.120484 | 0.0927389 | -1.299 | 0.193884 |
| cb.temperaturev4.l1 | 0.0056769 | 0.1081349 | 0.052 | 0.958132 |
| cb.temperaturev4.l2 | -0.0932972 | 0.0891686 | -1.046 | 0.295422 |
| cb.temperaturev4.l3 | -0.3942512 | 0.0948301 | -4.157 | <0.001 |
| cb.temperaturev4.l4 | 0.046028 | 0.0932205 | 0.494 | 0.62148 |
| cb.temperaturev5.l1 | 0.0921185 | 0.1746199 | 0.528 | 0.59782 |
| cb.temperaturev5.l2 | -0.0414536 | 0.1539405 | -0.269 | 0.787712 |
| cb.temperaturev5.l3 | -0.9669605 | 0.1395523 | -6.929 | <0.001 |
| cb.temperaturev5.l4 | -0.1748473 | 0.1354429 | -1.291 | 0.196728 |
| cb.temperaturev6.l1 | -0.0359648 | 0.2206378 | -0.163 | 0.870515 |
| cb.temperaturev6.l2 | 0.1178125 | 0.2095964 | 0.562 | 0.574053 |
| cb.temperaturev6.l3 | -0.5157761 | 0.1696462 | -3.04 | 0.002363 |
| cb.temperaturev6.l4 | -0.2525008 | 0.1463552 | -1.725 | 0.084481 |
| cb.humidityv1.l1 | -0.0760959 | 0.1415298 | -0.538 | 0.590807 |
| cb.humidityv1.l2 | 0.2974524 | 0.1564322 | 1.901 | 0.057239 |
| cb.humidityv1.l3 | 0.4239622 | 0.1249817 | 3.392 | <0.001 |
| cb.humidityv1.l4 | -0.3900315 | 0.1268123 | -3.076 | 0.0021 |
| cb.humidityv2.l1 | 0.2148096 | 0.095398 | 2.252 | 0.02434 |
| cb.humidityv2.l2 | 0.3397057 | 0.1095488 | 3.101 | 0.001929 |
| cb.humidityv2.l3 | 0.4692162 | 0.0831245 | 5.645 | <0.001 |
| cb.humidityv2.l4 | 0.0915356 | 0.0783017 | 1.169 | 0.242399 |
| cb.humidityv3.l1 | 0.0142524 | 0.114312 | 0.125 | 0.900777 |
| cb.humidityv3.l2 | 0.1509889 | 0.1272993 | 1.186 | 0.235585 |
| cb.humidityv3.l3 | 0.5810411 | 0.098065 | 5.925 | <0.001 |
| cb.humidityv3.l4 | -0.0502108 | 0.097828 | -0.513 | 0.607773 |
| cb.humidityv4.l1 | 0.2520897 | 0.1003122 | 2.513 | 0.011969 |
| cb.humidityv4.l2 | 0.3407618 | 0.1148536 | 2.967 | 0.003008 |
| cb.humidityv4.l3 | 0.5995064 | 0.0878938 | 6.821 | <0.001 |
| cb.humidityv4.l4 | -0.0240081 | 0.0868114 | -0.277 | 0.782122 |
| cb.humidityv5.l1 | -0.0465353 | 0.1176598 | -0.396 | 0.692469 |
| cb.humidityv5.l2 | 0.0943673 | 0.1310316 | 0.72 | 0.47141 |
| cb.humidityv5.l3 | 0.5635789 | 0.1014846 | 5.553 | <0.001 |
| cb.humidityv5.l4 | -0.0344524 | 0.0982732 | -0.351 | 0.725905 |
| cb.humidityv6.l1 | -0.0005845 | 0.1195719 | -0.005 | 0.9961 |
| cb.humidityv6.l2 | 0.3729215 | 0.1353168 | 2.756 | 0.005853 |
| cb.humidityv6.l3 | 0.2434892 | 0.1065491 | 2.285 | 0.022299 |
| cb.humidityv6.l4 | 0.1575367 | 0.1043284 | 1.51 | 0.131041 |

**Supplementary Table 6:** The estimated country-specific intercepts for each country for different selected months.

| Country | March to April | March to May | March to June | Full data set (i.e March to July) |
| --- | --- | --- | --- | --- |
| United Kingdom | -3.08±0.01 | -2.99±0.01 | -2.70±0.01 | -2.79±0.01 |
| Italy | -3.13±0.01 | -2.88±0.01 | -2.62±0.01 | -2.73±0.01 |
| France | -2.70±0.02 | -2.49±0.02 | -2.22±0.02 | -2.32±0.02 |
| Spain | -3.72±0.01 | -3.25±0.01 | -2.97±0.01 | -3.06±0.01 |
| Germany | -4.09±0.02 | -4.01±0.02 | -3.77±0.02 | -3.91±0.02 |
| Netherlands | -3.43±0.03 | -3.29±0.03 | -3.03±0.03 | -3.13±0.03 |
| Sweden | -3.86±0.05 | -3.97±0.04 | -3.83±0.03 | -4.00±0.03 |
| Romania | -3.19±0.09 | -3.45±0.06 | -3.26±0.05 | -3.43±0.04 |

**Supplementary Table 7:** The country-level sample size (number of days sampled) for the mixed model.

| Country | Sample size (day) |
| --- | --- |
| United Kingdom | 127 |
| Italy | 84 |
| France | 67 |
| Spain | 65 |
| Germany | 80 |
| Netherlands | 97 |
| Sweden | 129 |
| Romania | 125 |

